# Estimated SARS-CoV-2 Seroprevalence in Children and Adolescents in Mississippi, May Through September 2020

**DOI:** 10.1101/2021.02.05.21250792

**Authors:** Charlotte V. Hobbs, Jan Drobeniuc, Theresa Kittle, John Williams, Paul Byers, Subbian S. Panayampalli, Meagan Stephenson, Sara S. Kim, Manish M. Patel, Brendan Flannery, CDC COVID-19 Response Team, CDC COVID-19 Task Force Microbial Pathogenesis and Immune Response Laboratory.

## Abstract

Case-based tracking of COVID-19 in children and adolescents may underestimate infection, and compared with adults there is little pediatric SARS-CoV-2 seroprevalence data. To assess evidence of previous SARS-CoV-2 infections among children and adolescents in Mississippi, serologic testing for antibodies to SARS-CoV-2 was conducted on a convenience sample of residual serum specimens collected for routine laboratory testing by an academic medical center laboratory during May 17 through September 19, 2020. Seroprevalence by calendar month was standardized to the state population by race/ethnicity; cumulative numbers of infections were estimated by extrapolating seroprevalence to all those aged <18 years in Mississippi. Serum specimens from 1,603 individuals were tested; 175 (10.9%) were positive for SARS-CoV-2 antibodies. Among 1,579 (98.5%) individuals for whom race/ethnicity was known, the number testing positive was 16 (23.2%) of 69 Hispanic individuals, 117 (13.0%) of 901 non-Hispanic Black individuals and 30 (5.3%) of 565 non-Hispanic White individuals. Population-weighted seroprevalence estimates among those aged <18 years increased from 2.6% in May to 16.9% in September 2020. Cumulative numbers of infections extrapolated from seroprevalence data, however, were estimated at 117,805 (95% confidence interval [CI] = 68,771–168,708), suggesting that cases in children and adolescents are much higher than what was reported to the Mississippi State Department of Health (9,044 cases during this period). Further data to appreciate the burden of pediatric disease to inform public health policy is urgently needed.

## Introduction

As of January 2021, those aged <18 years accounted for >11% of reported coronavirus disease 2019 (COVID-19) cases in the United States^*^; however, data on pediatric infections with SARS-CoV-2, the virus that causes COVID-19, are limited (*1*). Surveys of SARS-CoV-2 antibody seroprevalence suggest that cumulative incidence of infection is much higher than that ascertained by reported COVID-19 cases (*2,3*), and further data to appreciate the burden of pediatric disease to inform public health policy is urgently needed.

Most persons who are infected with SARS-CoV-2 develop antibodies to SARS-CoV-2 proteins within 1–2 weeks of disease onset (*4*). Serologic testing for SARS-CoV-2 antibodies, albeit having imperfect sensitivity and specificity,^†^ is useful to identify past SARS-CoV-2 infections. Serology tests are being used widely in seroprevalence studies to understand patterns of virus spread and cumulative incidence of SARS-CoV-2 infection (*2,3*).^§^

To assess evidence of previous SARS-CoV-2 infections among children and adolescents in Mississippi during early months of the COVID-19 pandemic, we conducted serologic testing for antibodies to SARS-CoV-2 in a convenience sample of residual serum specimens collected for routine clinical or diagnostic testing from May–September, 2020. We projected monthly seroprevalence to estimate cumulative SARS-CoV-2 infections over time and compared projections with cumulative numbers of Mississippi State Department of Health (MSDH)-reported COVID-19 cases among children and adolescents in Mississippi.

## Methods

This retrospective seroprevalence study was conducted by the University of Mississippi Medical Center to describe trends in SARS-CoV-2 antibody seroprevalence among children and adolescents in Mississippi during the COVID-19 pandemic. The University of Mississippi Medical Center provides clinical laboratory services for university hospitals in central Mississippi and 12 hospitals outside the university network statewide (*5*). Demographic data including age, sex, race/ethnicity and date of collection were obtained for deidentified residual serum specimens collected for routine clinical testing during May 17–September 19, 2020 from those aged <18 years. One specimen per person was included in the analysis, either the first seropositive specimen or the earliest specimen from individuals with all seronegative specimens to avoid potential bias in underestimating infections from decline in antibodies below the limit of detection for seropositivity. Sera were stored at –20°C (–4°F) before testing at CDC.

Seropositivity was determined for serum specimens using one of two assays, based on specimen volume. Specimens with adequate volume (≥ 0.3 mL) were tested using a qualitative VITROS anti–SARS-CoV-2 total antibody in vitro diagnostic test using the automated VITROS 3600 Immunodiagnostic System (Ortho Clinical Diagnostics) (*6*). Briefly, one aliquot was heat-treated at 56°C (132.8°F) for 10 minutes and tested on the VITROS Immunodiagnostic System. An automatically calculated ratio of test sample signal to cutoff value (S/C) <1.0 was interpreted as nonreactive and S/C ≥ 1.0 was interpreted as reactive for anti–SARS-CoV-2 total antibody (*6*). Samples with volumes <0.3 mL were tested to determine seropositivity using an enzyme linked immunosorbent assay (ELISA) developed by CDC to measure total SARS-CoV-2 antibodies against the extracellular domain of the SARS-CoV-2 spike protein (*2*). Both assays were granted Food and Drug Administration Emergency Use Authorization.

Seroprevalence by calendar month was standardized to the Mississippi population aged <18 years by race/ethnicity^**^; 95% CIs accounting for assay test performance were estimated using published methods (*2*). Cumulative numbers of SARS-CoV-2 infections, estimated by extrapolating seroprevalence and 95% CIs to the Mississippi population aged <18 years were compared with cumulative numbers of confirmed and probable COVID-19 cases (as defined by the Council of State and Territorial Epidemiologists)^††^ in those aged <18 years reported to the Mississippi State Department of Health.^§§^ Ratios of estimated SARS-CoV-2 infections to reported COVID-19 cases were calculated by dividing estimated numbers of SARS-CoV-2 infections by the reported cumulative number of COVID-19 cases as of the last day of the preceding month. Statistical analyses were conducted using SAS (version 9.4; SAS Institute). This activity was reviewed by CDC and was conducted consistent with applicable federal law and CDC policy. ^¶¶^ The project was also reviewed and approved by the University of Mississippi Medical Center Institutional Review Board through the expedited review procedure.

## Results

Among 1,603 individuals aged <18 years included in analyses, serum specimens from 175 (10.9%) tested positive for SARS-CoV-2 antibodies [10.4% (152 of 1469) by VITROS assay and 17.2% (23 of 134) by ELISA assay] (Table 1). After adjusting by race/ethnicity to the Mississippi population aged <18 years, population-weighted seroprevalence estimates seroprevalence increased from 2.6% in May to 16.9% in September and paralleled MSDH reported case data (Figure). Extrapolating to the state population, an estimated 117,805 (95% CI = 68,771–168,708) Mississippi residents aged <18 years might have been infected with SARS-CoV-2 by mid-September, 2020; through August 31, a total of 9,044 COVID-19 cases among those aged <18 years had been reported to MSDH (Table 2).

**Figure 1.**
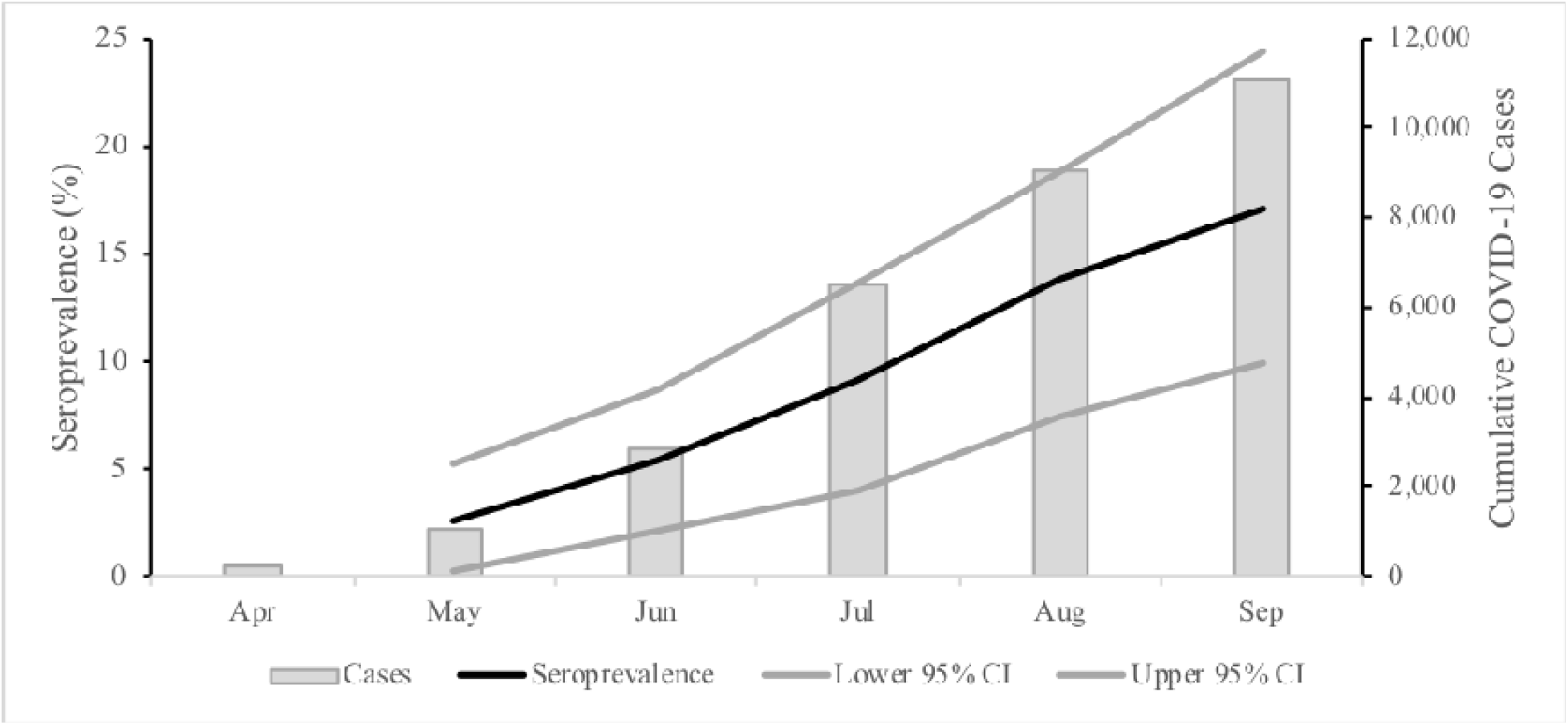
Estimated race/ethnicity–standardized SARS-CoV-2 antibody seroprevalence*and cumulative number of reported COVID-19 cases aged <18 years — Mississippi, April– September 2020. * from residual serum specimens collected May 17–September 19, 2020, from individuals aged <18 years.

**Table 1.**
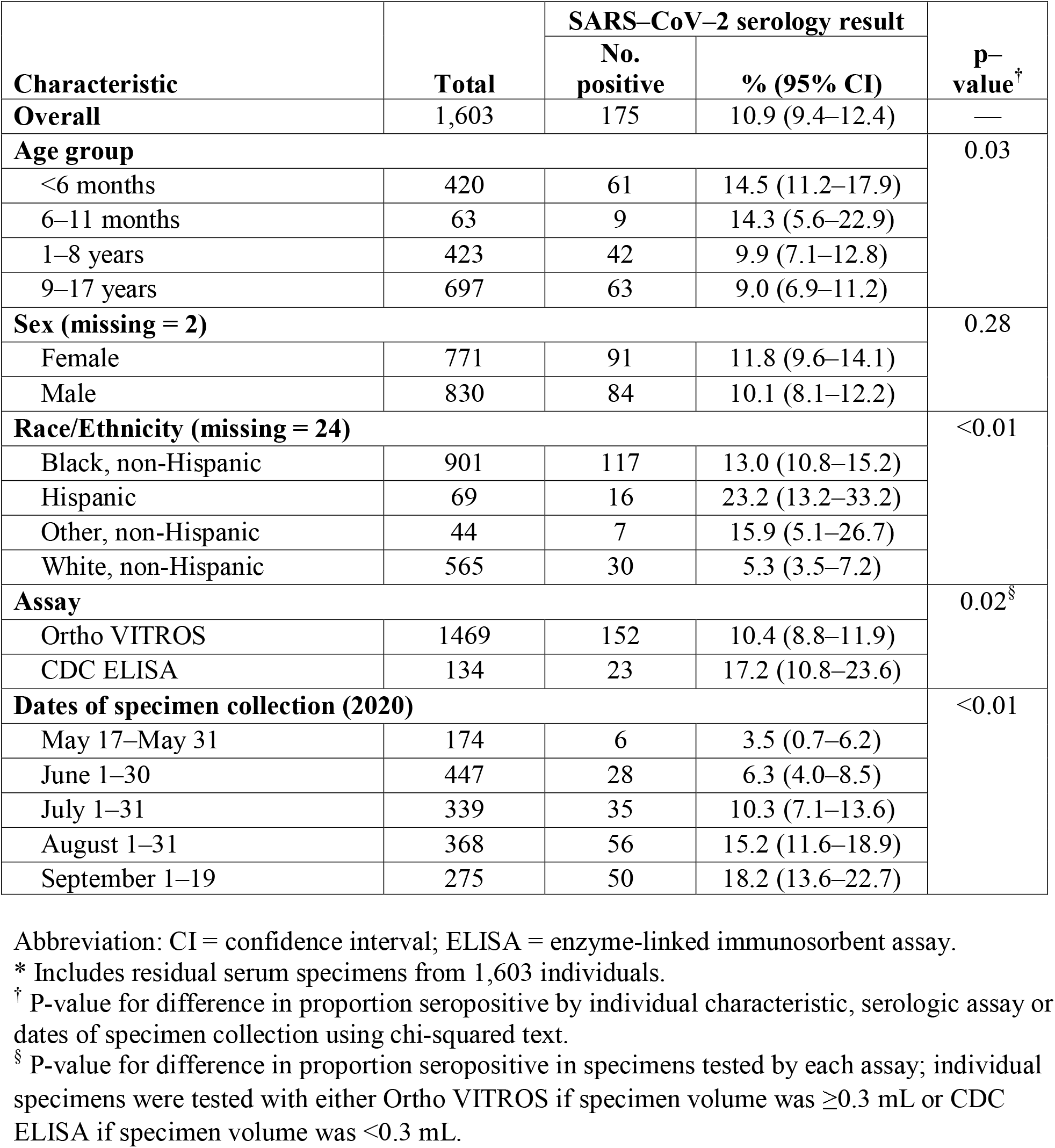
Characteristics and serology results of Children and Adolescents aged <18 years with serum specimens* tested for presence of SARS-CoV-2 antibodies — Mississippi, May 17—September 19, 2020.

| Characteristic | Total | SARS–CoV–2 serology result |  | p-value<sup>†</sup> |
| --- | --- | --- | --- | --- |
|  |  | No. positive | % (95% CI) |  |
| **Overall** | 1,603 | 175 | 10.9 (9.4–12.4) | — |
| **Age group** |  |  |  | 0.03 |
| <6 months | 420 | 61 | 14.5 (11.2–17.9) |  |
| 6–11 months | 63 | 9 | 14.3 (5.6–22.9) |  |
| 1–8 years | 423 | 42 | 9.9 (7.1–12.8) |  |
| 9–17 years | 697 | 63 | 9.0 (6.9–11.2) |  |
| **Sex (missing = 2)** |  |  |  | 0.28 |
| Female | 771 | 91 | 11.8 (9.6–14.1) |  |
| Male | 830 | 84 | 10.1 (8.1–12.2) |  |
| **Race/Ethnicity (missing = 24)** |  |  |  | <0.01 |
| Black, non-Hispanic | 901 | 117 | 13.0 (10.8–15.2) |  |
| Hispanic | 69 | 16 | 23.2 (13.2–33.2) |  |
| Other, non-Hispanic | 44 | 7 | 15.9 (5.1–26.7) |  |
| White, non-Hispanic | 565 | 30 | 5.3 (3.5–7.2) |  |
| **Assay** |  |  |  | 0.02<sup>§</sup> |
| Ortho VITROS | 1469 | 152 | 10.4 (8.8–11.9) |  |
| CDC ELISA | 134 | 23 | 17.2 (10.8–23.6) |  |
| **Dates of specimen collection (2020)** |  |  |  | <0.01 |
| May 17–May 31 | 174 | 6 | 3.5 (0.7–6.2) |  |
| June 1–30 | 447 | 28 | 6.3 (4.0–8.5) |  |
| July 1–31 | 339 | 35 | 10.3 (7.1–13.6) |  |
| August 1–31 | 368 | 56 | 15.2 (11.6–18.9) |  |
| September 1–19 | 275 | 50 | 18.2 (13.6–22.7) |  |

Abbreviation: CI = confidence interval; ELISA = enzyme-linked immunosorbent assay.
\* Includes residual serum specimens from 1,603 individuals.
<sup>†</sup> P-value for difference in proportion seropositive by individual characteristic, serologic assay or dates of specimen collection using chi-squared test.
<sup>§</sup> P-value for difference in proportion seropositive in specimens tested by each assay; individual specimens were tested with either Ortho VITROS if specimen volume was ≥0.3 mL or CDC ELISA if specimen volume was <0.3 mL.

**Table 2.** Estimated number of SARS-CoV-2 infections based on seroprevalence estimates and the number of reported COVID-19 cases at of the end of the preceding month for Children and Adolescents aged <18 years — Mississippi, May 17–September 19, 2020.

| **Date of serum specimen collection** | **Estimated seroprevalence\***<br>**% (95% CI)** | **Estimated cumulative infections<sup>†</sup> No. (95% CI)** | **Cases reported<sup>§</sup>**<br>**(Report date)** | **Estimated infections/reported cases (95% CI)** |
| --- | --- | --- | --- | --- |
| May 17–31 | 2.6 (0.3–5.2) | 17,825 (2,092–36,316) | 256 (Apr 30) | 69.6 (8.2–141.9) |
| June 1–30 | 5.4 (2.2–8.6) | 37,336 (15,097–59,905) | 1,087 (May 31) | 34.4 (13.9–55.1) |
| July 1–31 | 8.9 (3.8–13.3) | 62,092 (26,689–93,023) | 2,881 (Jun 30) | 21.6 (9.3–32.3) |
| August 1–31 | 13.5 (7.3–18.5) | 94,179 (50,943–129,208) | 6,476 (Jul 31) | 14.5 (7.9–20.0) |
| September 1–19 | 16.9 (9.9–24.2) | 117,805 (68,771–168,708) | 9,044 (Aug 31) | 13.0 (7.6–18.7) |

Abbreviation: CI = confidence interval.
\* Standardized to the race/ethnicity distribution of the Mississippi population aged <18 years using weights derived from the 2019 US census; 95% confidence intervals for seroprevalence by month were calculated assuming a binomial distribution for SARS-CoV-2 seropositivity in the sample standardized for race/ethnicity.
<sup>†</sup> Race/ethnicity-standardized seroprevalence estimates for each time period applied to the 2019 Mississippi population aged <18 years = 698,420 (www.census.gov).
<sup>§</sup> Cumulative number of confirmed and probable COVID-19 cases in those aged <18 years reported to Mississippi State Department of Health as of the end of the preceding month (source: Mississippi State Department of Health).

Among 1,579 individuals with known race/ethnicity, 16 (23.2%) of 69 Hispanic individuals, 117 (13.0%) of 901 non-Hispanic Black individuals and 30 (5.3%) of 565 non-Hispanic White individuals tested seropositive.

Ratios of estimated numbers of SARS-CoV-2 cases based on seropositivity to COVID-19 cases based on reporting in each calendar month decreased from 69.6 (95% CI = 8.2–141.9) in May to 13.0 (95% CI = 7.6–18.7) in September.

## Discussion

In analyses of a convenience sample of residual sera collected from those <18 years old from May-September 2020 in Mississippi, 18.2% of tested individuals aged <18 years showed evidence of previous SARS-CoV-2 infection (16.9% by population-weighted seroprevalence estimates). Monthly increases in population-weighted seroprevalence paralleled increases in MSDH reported COVID-19 cases. However, projected cumulative infections based on seroprevalence suggests that case-based surveillance significantly underestimated SARS-CoV-2 infections among children and adolescents. This is consistent with national estimates of COVID-19 disease incidence in all ages (*7*).***

Significantly, seropositivity among non-Hispanic Black and Hispanic individuals was 2.4 and 4.3 times higher, respectively, than that among non-Hispanic White individuals. These data reflect previously published data in which substantially higher positivity rates for SARS-CoV-2 testing were noted in minority individuals compared with white non-minority individuals in Mississippi (*5*). Also, the noted decrease of SARS-CoV-2 cases based on seropositivity to COVID-19 cases based on reporting in each calendar month suggests an improvement in case detection over time, even though the number of SARS-CoV-2 cases estimated based on seroprevalence was consistently higher than the number of SARS-CoV-2 cases reported during each month.

Nationwide serosurveys have identified varying seroprevalences by sex, age group, and urban/rural status. In four cross-sectional serosurveys conducted in Mississippi during July– October 2020, female sex, age 18–49 years, and living in nonmetropolitan jurisdictions were associated with higher SARS-CoV-2 seroprevalence (*3*). However, the number of specimens from those aged <18 years were insufficient to provide seroprevalence estimates for this age group. In contrast, the current investigation of seropositivity benefited from large numbers of pediatric specimens collected during a 4-month period when incidence of reported COVID-19 cases increased rapidly.

It is also of note that compared with seroprevalence data from older age groups in Mississippi, data from this sample suggested that cumulative infection rates by mid-September among individuals aged <18 years were similar to those among individuals aged 18–49 years, the age group with the highest seroprevalence during the period (*3*).

The findings in this report are subject to at least four limitations. First, seropositivity among a convenience sample of sera from one laboratory might not be representative of seropositivity among individuals aged <18 years in Mississippi; therefore, comparisons between these estimates of cumulative SARS-CoV-2 infections and reported COVID-19 cases in Mississippi should be interpreted with caution. Second, individuals who have blood collected for routine laboratory testing might differ from the general pediatric population in underlying health conditions, access to care or adherence to prevention measures, including use of masks and social distancing. However, compared with more representative serosurveys, residual sera from commercial laboratories have previously been shown to provide an approximate measure of community seroprevalence (*3*). Third, misclassification of antibody status was possible due to possible imperfect sensitivity and specificity of the assays used in the report. Finally, selecting the first seropositive specimen from individuals testing positive at any time point rather than randomly selected specimens might have overestimated population seroprevalence.

Alternatively, seroprevalence could be underestimated if participants who were infected had not yet mounted an antibody response or if antibody titers had declined since infection (*8,9*), and it is also noted that positive antibody status in the infant group could reflect SARS-CoV-2 immunoglobulin G placental transfer from an infected mother.

In conclusion, these SARS-CoV-2 infection estimates among children and adolescents in Mississippi add to those from other studies of the general population from nationwide cross-sectional serosurveys. The higher infection rate estimated from seroprevalence data compared with reported cases suggests disease burden in children and adolescents is much greater than appreciated, with a predominance of infections in minority individuals. The comparable seroprevalence in children and adolescents compared with adults observed in this dataset may be underappreciated, as other studies have estimated that children and adolescents have estimated lower susceptibility to SARS-CoV-2 infection compared with adults (*10*). These data also highlight the importance of COVID-19 mitigation measures, such as masking and social distancing, to protect children and adolescents from infection. Further such studies are urgently needed to characterize COVID-19 pediatric disease burden and, in turn, inform public health measures for children and adolescents.

## Data Availability

All relevant data are included in the manuscript.

## Acknowledgments

Lora M. Martin, Children’s of Mississippi and School of Nursing, University of Mississippi Medical Center, Jackson, Mississippi; Eugene Melvin, Center for Informatics, University of Mississippi Medical Center, Jackson, Mississippi.

## Disclaimer

The findings and conclusions in this report are those of the authors and do not necessarily represent the official position of the Centers for Disease Control and Prevention.

## Footnotes

* https://covid.cdc.gov/covid-data-tracker/#demographics

† https://www.cdc.gov/coronavirus/2019-ncov/lab/resources/antibody-tests-guidelines.html.

§ https://covid.cdc.gov/covid-data-tracker/#national-lab.

¶ https://www.biorxiv.org/content/10.1101/2020.04.24.057323v2.

** 2019 US census data for Mississippi, accessed at https://wonder.cdc.gov.

†† https://wwwn.cdc.gov/nndss/conditions/coronavirus-disease-2019-covid-19/case-definition/2020/

§§ https://msdh.ms.gov/msdhsite/_static/14,0,420.html

¶¶ Activity was determined to meet the requirements of public health surveillance as defined in 45 CFR 46.102(l)(2).

*** https://www.cdc.gov/coronavirus/2019-ncov/cases-updates/burden.html.

## Notes

### Competing Interest Statement

The authors have declared no competing interest.

### Clinical Trial

de-identified samples analyzed only

### Author Declarations

This activity was reviewed by CDC and was conducted consistent with applicable federal law and CDC policy (Activity was determined to meet the requirements of public health surveillance as defined in 45 CFR 46.102(l)(2).) The project was also reviewed and approved by the University of Mississippi Medical Center Institutional Review Board through the expedited review procedure.

